# Effects of interval treadmill training on spatiotemporal parameters in children with cerebral palsy: a machine learning approach

**DOI:** 10.1101/2023.12.20.23300157

**Authors:** Charlotte D. Caskey, Siddhi R. Shrivastav, Alyssa M. Spomer, Kristie F. Bjornson, Desiree Roge, Chet T. Moritz, Katherine M. Steele

## Abstract

**Background:** Children with cerebral palsy (CP) have reduced step length, reduced symmetry, and greater step width compared to their peers. Short-burst interval locomotor treadmill training (SBLTT) is a novel rehabilitation paradigm for children with CP that may improve spatiotemporal outcomes. However, for interventions like SBLTT, quantifying rehabilitation responses and optimizing therapy parameters remains challenging. Machine learning and causal modeling provide a platform to quantify step-by-step changes during gait training to understand mechanisms driving individual responses.

**Research question:** What is the direct effect of SBLTT on step length, asymmetry, and step width in children with CP?

**Methods:** We recruited four children with spastic CP, ages 4-13. Each participant received 24 sessions of SBLTT over 8-12 weeks, with spatiotemporal outcomes monitored with an instrumented treadmill. We used Bayesian Additive Regression Trees (BART) to model the direct effect of therapy parameters on step length, step length asymmetry, and step width. Additionally, we generated *in silico* data for 150 virtual participants to quantify the quality of BART models to capture rehabilitation progression.

**Results:** After SBLTT, participants’ step lengths increased by 26 ± 13% (pre-post effect). Controlling for treadmill speed, time in session, limb, and treadmill incline with BART demonstrated that SBLTT directly increased step length for three participants (direct effect: 13.5 ± 4.5%), while one participant decreased step length (−11.6%). SBLTT had minimal effects on step length asymmetry and step width. Virtual datasets demonstrated that BART could accurately predict step length progression (R^2^ = 0.73) and plateaus in progression (R^2^ = 0.87), with better model fit for participants with less step-to-step variability.

**Significance:** Tools such as BART can leverage step-by-step data collected during gait training to monitor progression and optimize rehabilitation protocols. This work can help personalize rehabilitation and understand the causal mechanisms driving individual responses.

## Introduction

Daily walking requires the capacity to quickly modulate speed, step length, and step width to navigate natural environments. Speed and stability modulation are especially important at a young age, as children frequently transition between low and high intensity activities during play [1–3]. For children with cerebral palsy (CP), reductions in physical activity result in fewer opportunities to practice speed modulation, hindering development of robust and flexible movement patterns [4–7]. As such, gait training for ambulatory children with CP often focuses on increasing walking speed, step length, and stability. Short-burst interval locomotor treadmill training (SBLTT) was recently developed specifically for children with CP and has been shown to improve walking speed, endurance, and community mobility for children with CP [8]. However, it is unknown whether SBLTT impacts children’s speed modulation strategies or whether it leads to improvements in step length, symmetry, and step width.

A fundamental challenge in gait rehabilitation is quantifying the direct effect of an intervention on targeted outcomes. Training programs like SBLTT have a range of individualizable parameters that vary across sessions, such as duration of training, treadmill speed, and treadmill incline. This makes isolating the effect of training on a specific outcome challenging. Causal inference and machine learning help address these limitations by quantifying the direct effect of individualizable parameters on rehabilitation responses. Specifically, Directed Acyclic Graphs (DAGs) can be used to graphically model the assumed relationship between study variables. Traditional results, or pre-post effects, consider all direct and indirect (i.e., mediated) relationships between a variable and the outcome using only the first and last session. Alternatively, a DAG can help determine the adjustment set needed to quantify direct effects, controlling for all intermediaries that may bias the outcome. The identified adjustment set can then be input into a statistical model to understand how each variable contributes to the observed response. Machine learning methods like Bayesian Additive Regression Trees (BART) have emerged as favorable statistical model for handling the nonlinear, interacting effects inherent in gait [9–11]. BART is a “sum of trees” regression method capable of modeling relationships between input and response variables with low levels of bias and variance [12]. Prior research has leveraged both DAGs and BART to understand complex factors influencing biomechanics and treatment response in CP [9,13–15]. However, BART modeling informed by DAGs has not been used to model individual treatment responses over time.

The purpose of this study was to quantify the pre-post and direct effects of 24 sessions of SBLTT on speed modulation in four children with CP. We hypothesized that SBLTT would have a direct effect on increasing step length, decreasing step length asymmetry, and decreasing step width for children with CP. Secondarily, we leveraged data from these four participants to generate *in silico* datasets to quantify BART accuracy for monitoring rehabilitation responses. We hypothesized that BART would be able to accurately predict rehabilitation responses in the virtual dataset, including correctly identifying the rate in which step length changes between sessions and identifying plateau points.

## Methods

### Participants

We recorded spatiotemporal data for four children with CP who participated in 24 sessions of SBLTT (Table 1). Our inclusion criteria were children aged 4-17 years and Gross Motor Functional Classification System (GMFCS) Levels I-II [16], who could walk 20 yards and could follow simple cued motor tasks. We excluded participants who had prior selective dorsal rhizotomy, botulinum toxin injections in the last six months, orthopedic surgery in the last 12 months, or a history of uncontrolled seizures. All caregivers and participants provided written informed consent and age-appropriate assent. This study was approved by the University of Washington Institutional Review Board (STUDY00008896) and was registered as ClinicalTrials.gov #NCT04467437.

**Table 1.** Participant Characteristics.

| Participant Identifier | Sex | Age Range (years) | Leg Length (m) | Diagnosis | More-affected side | GMFCS Level | Assistive Devices |
| --- | --- | --- | --- | --- | --- | --- | --- |
| P01 | M | 8-13 | 0.81 | Spastic diplegia | Left | II | Bilateral AFO-FC |
| P02 | M | 2-7 | 0.50 | Spastic diplegia | Left | I | Bilateral AFO-FC |
| P03 | M | 8-13 | 0.75 | Spastic diplegia | Right | II | Bilateral AFO-FC |
| P04 | M | 8-13 | 0.85 | Spastic hemiplegia | Left | I | Left lift |

The more-affected side is based on the side with more spasticity at baseline and parent reports. GMFCS = Gross Motor Function Classification System; AFO-FC = ankle foot orthoses footwear combination

Each training session included a 5–15-minute active warm-up either overground or on the treadmill, 30-minutes of SBLTT, and a 5-minute active cool-down. Rest breaks were provided as needed, and participants were encouraged to use handrails for safety. SBLTT consisted of 30-second bursts, alternating between walking at a slow and a fast speed to mimic natural variability children experience during walking in daily life. The initial slow and fast starting speeds were selected based on each individual’s self-selected and fast 10-meter overground walking speeds, respectively [8]. The slow speed was kept constant across all sessions, while the fast speed was increased based on self-reported and clinician-perceived exertion per a predetermined protocol during and across sessions [17]. If the speed was increased to the point that the participant began to run, the incline of the treadmill was increased in 1% increments until walking was maintained, which occurred for P01 and P04. During training, all participants wore the orthoses that they used during community walking, including ankle foot orthoses footwear combination (AFO-FC) and shoe lifts (Table 1).

To evaluate step-by-step responses, plantar pressure data were collected during five of the SBLTT training sessions using a pressure-instrumented treadmill (h/p/cosmos, Zebris Medical GmbH, Isny, Germany; Figure 1A). Pressure data were collected at 300 Hz through the Noraxon MyoPressure Software MR3 (Noraxon U.S.A., Inc., Scottsdale, AZ, USA). MR3 performed automatic step detection, which was confirmed by a researcher. We focused our analyses on three spatiotemporal outcomes: step length, step length asymmetry, and step width. Only the fast speed bursts were used in the analysis as the fast speeds progressed during training to maintain an appropriate exertion level, making them the primary point of interest. Step length was defined as the anterior-posterior distance between the center of pressure at the initial point of contact of subsequent foot strikes. Step length asymmetry was calculated using the asymmetry index (ASI) and represented as a percent difference of consecutive steps [18]:

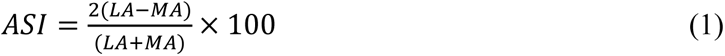

**Figure 1.**
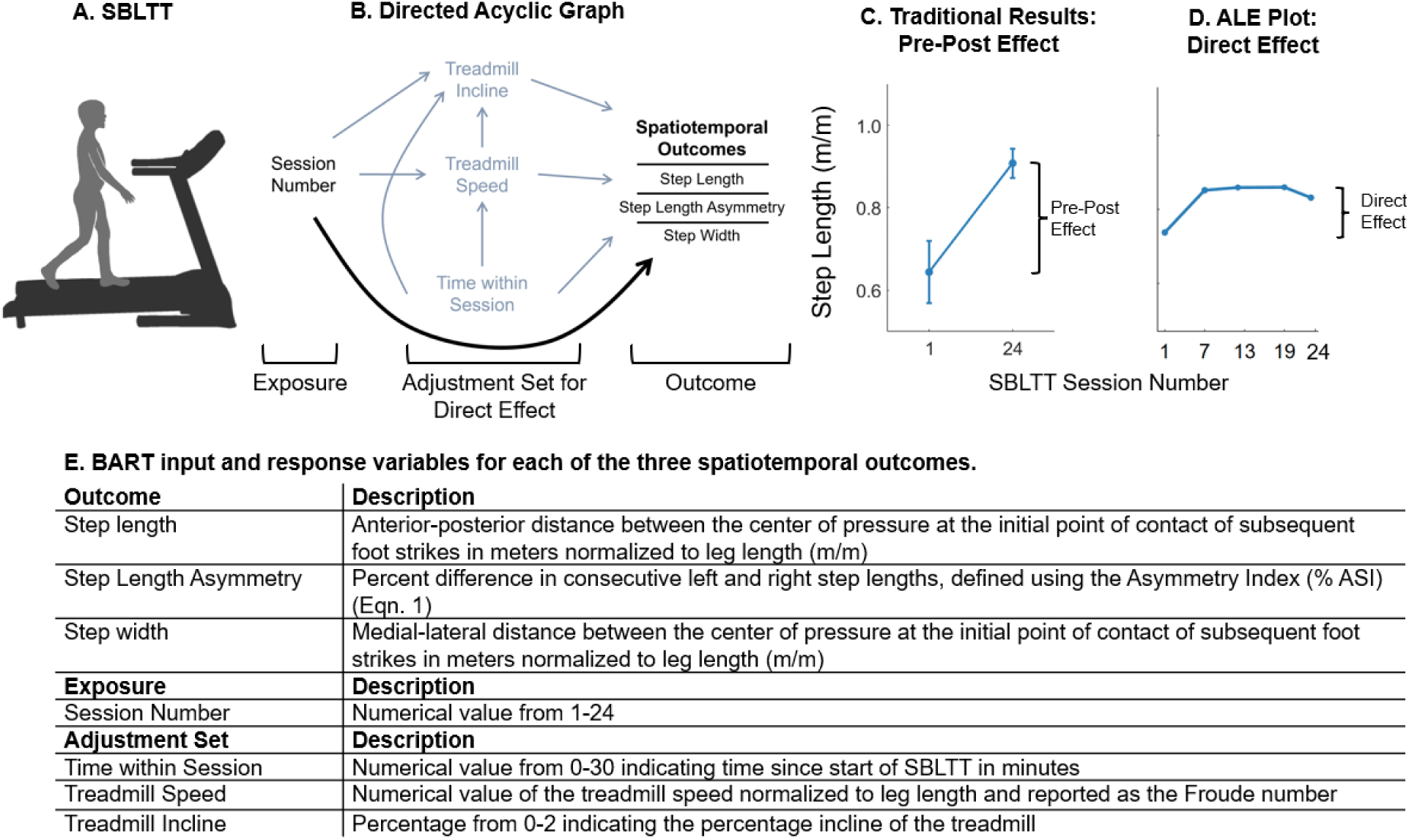
A) Step length and width were recorded during sessions of Short-burst Interval Locomotor Treadmill Training (SBLTT) using a pressure-instrumented treadmill. B) A Directed Acyclic Graph (DAG) describing the assumed causal relationship between session number (exposure) and spatiotemporal parameters (outcomes). Spatiotemporal outcomes included step length, step length asymmetry, and step width. The adjustment set for direct effects (black arrow) includes time in session, treadmill speed, treadmill incline, and side of the body. No adjustment set is needed when calculating pre-post effects. C) The pre-post effect of SBLTT on spatiotemporal outcomes were calculated using the first and lasts training visit. D) A Bayesian Additive Regression Tree (BART) model was generated for each participant and outcome and used to quantify the direct effect of the input parameters on each spatiotemporal outcome. The direct effect was calculated as the change in outcome from Accumulated Local Effects (ALE) plot for each model input. E) BART input and response variables for each of the spatiotemporal models.

where the less-affected (LA) and more-affected (MA) legs were defined as the sides with less or more spasticity at baseline, respectively, as determined by physical exam and parent reports. Step width was defined as the medial-lateral distance between the center of pressure at the initial point of contact of subsequent foot strikes. All spatiotemporal parameters were calculated with custom scripts in MATLAB (Mathworks, Natick, MA, USA) from the MR3 center of pressure at initial contact for all left and right steps. Step length, step width, and speed were normalized to each participant’s leg length, with speed reported as the Froude number [19].

### Modeling Framework

We built a DAG based on the individualizable parameters during SBLTT to identify the adjustment set needed to quantify direct effects of SBLTT on gait including: treadmill speed, incline, time within session, and side of the body (Figure 1B). The pre-post effect was calculated using the first and last training visit and did not require an adjustment set, as this is most commonly reported in the rehabilitation literature (Figure 1C). We defined the number of SBLTT training sessions as the exposure and the three spatiotemporal parameters (step length, step length asymmetry, step width) as individual outcome variables for three separate BART models per participant.

BART models were created for each spatiotemporal outcome and each participant using the SBLTT fast speed bursts (Figure 1E). The BART models were generated using 10-fold cross-validation for hyperparameter tuning in RStudio (R Version 4.1.3; *bartMachine* package) [20]. For replicability, we set the seed of each model to 18. R^2^ is reported throughout as a measure of model fit, with an R^2^ = 1 indicating the BART model was able to fully capture variability in the dataset with given information.

Accumulated local effect (ALE) plots were used to evaluate the direct effect of each input variable on each spatiotemporal outcome to illustrate how training parameters influence walking function [21]. For each input variable, the ALE plots were created using a bootstrapping procedure (n=3), wherein a sample of 75% of the total data were used to generate a series of ALE plots from which the average (± standard deviation) trend was generated (*ALEPlot* package) [22]. These data were binned into evenly space increments as a smoothing procedure. The direct effect of each input variable on the response variable was quantified as the absolute difference between the 10^th^-90^th^ percentile of the outcome for each ALE plot [14] (Figure 1D).

### In Silico Model Validation

To evaluate the accuracy of BART in quantifying changes in spatiotemporal outcomes across sessions, we also generated virtual datasets to supplement our experimental data using custom MATLAB scripts. Each virtual participant was randomly assigned an initial walking speed, step length, step length variability, and rate at which step length increased with training progression based on the data from our four participants. To evaluate prediction accuracy of BART, we fit a linear model to the predicted rate of change, calculated from the average ALE plot slope and compared against the specified rate of change. We report the linear fit (R^2^) between predicted and specified values as a measure of model accuracy which converged by 150 participants.

To evaluate the impact of step-to-step variability on BART model fit, we used the same virtual participants described above, but changed the standard deviation of the variability of each virtual participant’s step length, ranging 1-45 millimeters (mm). We used linear regression to evaluate whether step-to-step variability was associated with BART model fit (R^2^). Similarly, we modeled the relationship between each virtual participant’s specified variability and the error of the predicted step length progression using linear regression to quantify the relationship between step-to-step variability and BART model accuracy. To measure the accuracy of BART to detect plateaus in an individual’s rehabilitation progression, we randomly specified a plateau at the 1st, 6th, 12th, 18th, or 24th visit, while the relationship between step length and other variables were unchanged. We compared the association between the specified and predicted plateau points (ALE plot slope less than 0.6 mm/session) using linear regression.

## Results

### Impact of SBLTT on Step Length

SBLTT had a positive pre-post effect on step length, as participants increased step length during fast speed bursts by 26 ± 13% from the first to last SBLTT session (Figure 2A). This was paralleled by a 55 ± 24 % increase in treadmill speed (Froude number: +0.20 ± 0.067). Evaluating the direct effects of SBLTT on step length with BART indicated that three participants increased step length by an average of 13.5 ± 4.5% after controlling for all other variables in the DAG (direct effect: 0.11 ± 0.035 m/m; Figure 2B and D). The individualized BART models predicted step length with high model fit (0.77 < R^2^ < 0.94) for these three participants (Figure 2C). For the remaining youngest participant (P02), step length decreased by 11.4% (direct effect: 0.098 m/m). However, BART model fit was also much lower for P02 (R^2^ = 0.21), potentially reflecting P02’s greater step-to-step variability (P01: 19 mm; P02: 62 mm; P03: 30 mm; P04: 42 mm).

**Figure 2.**
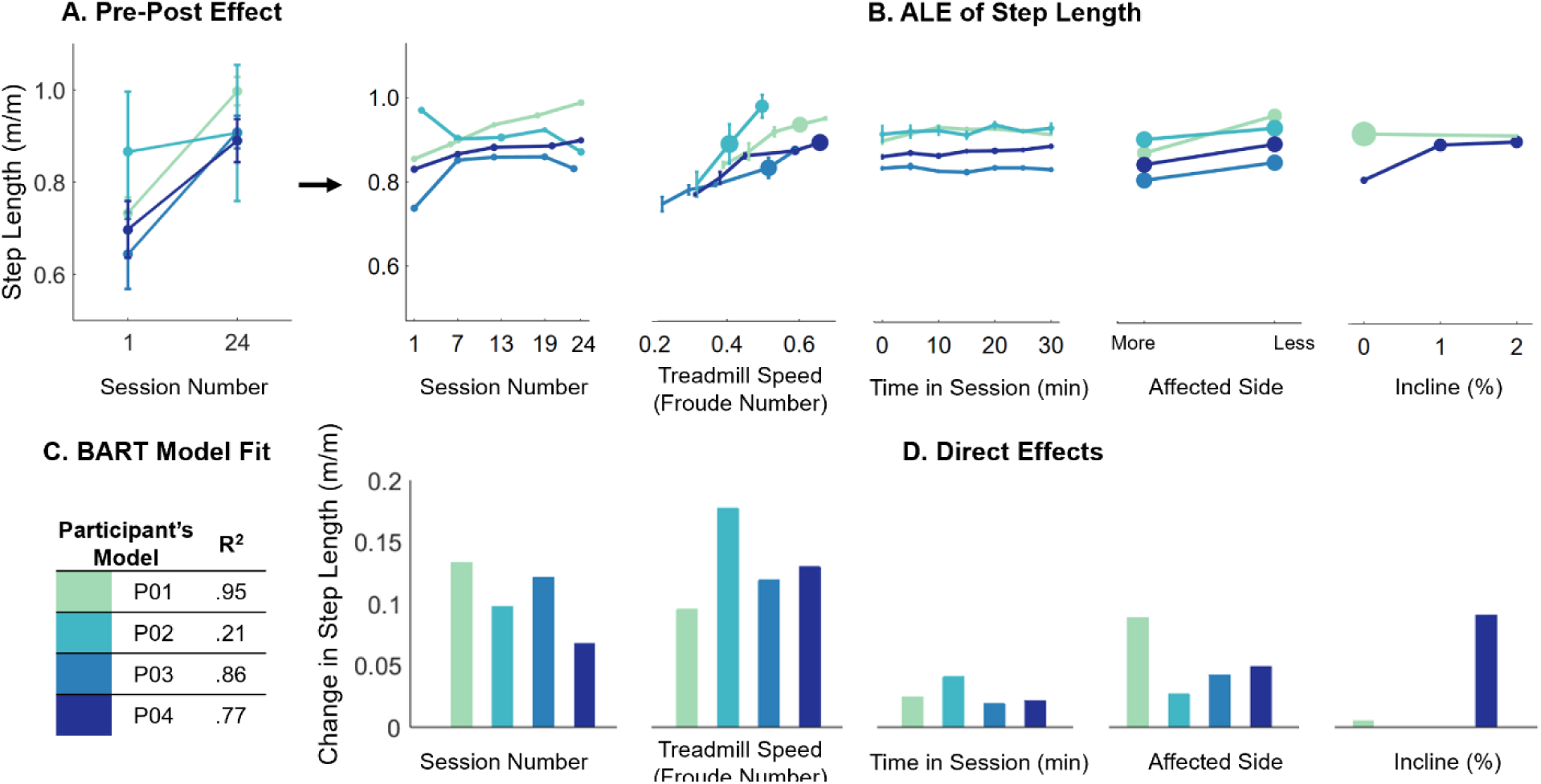
A) Pre-post effect of SBLTT on step length for the more affected side. B) BART results quantify direct effects of SBLTT on step length. Accumulated Local Effects (ALE) plots for each input variable show the effect of that variable on step length including session number, treadmill speed (Froude number), time within session, side, and treadmill incline. The size of the data point on each ALE plot depicts the relative number of data points in each bin. C) BART model fit (R^2^) for each participant. D) Direct effects of each input variable on the response variable, step length, calculated from the change in the ALE plots in B).

### Impact of SBLTT on Step Length Asymmetry

The pre-post and direct effects of SBLTT on step length Asymmetry Index (ASI) were minimal. The pre-post effects indicated that ASI increased by 2.1 ± 4.3 percentage points with SBLTT during the fast speed bursts (Figure 3A). All participants favored a longer step on the less-affected side. The BART models had poor to moderate fit for ASI (0.08 < R^2^ < 0.64; Figure 3C). SBLTT session number had a direct effect of decreasing ASI for two participants (P01: 5.6 percentage points; P04: 10 percentage points) and increasing ASI for two participants (P02: 5.5 percentage points; P03: 4.5 percentage points; Figure 3B and D). Time within session had a small effect on ASI for the diplegic participants (direct effect < 3.0 percentage points), but the hemiplegic participant (P04) consistently became more asymmetric during the time in session (direct effect = 5.2 percentage points).

**Figure 3.**
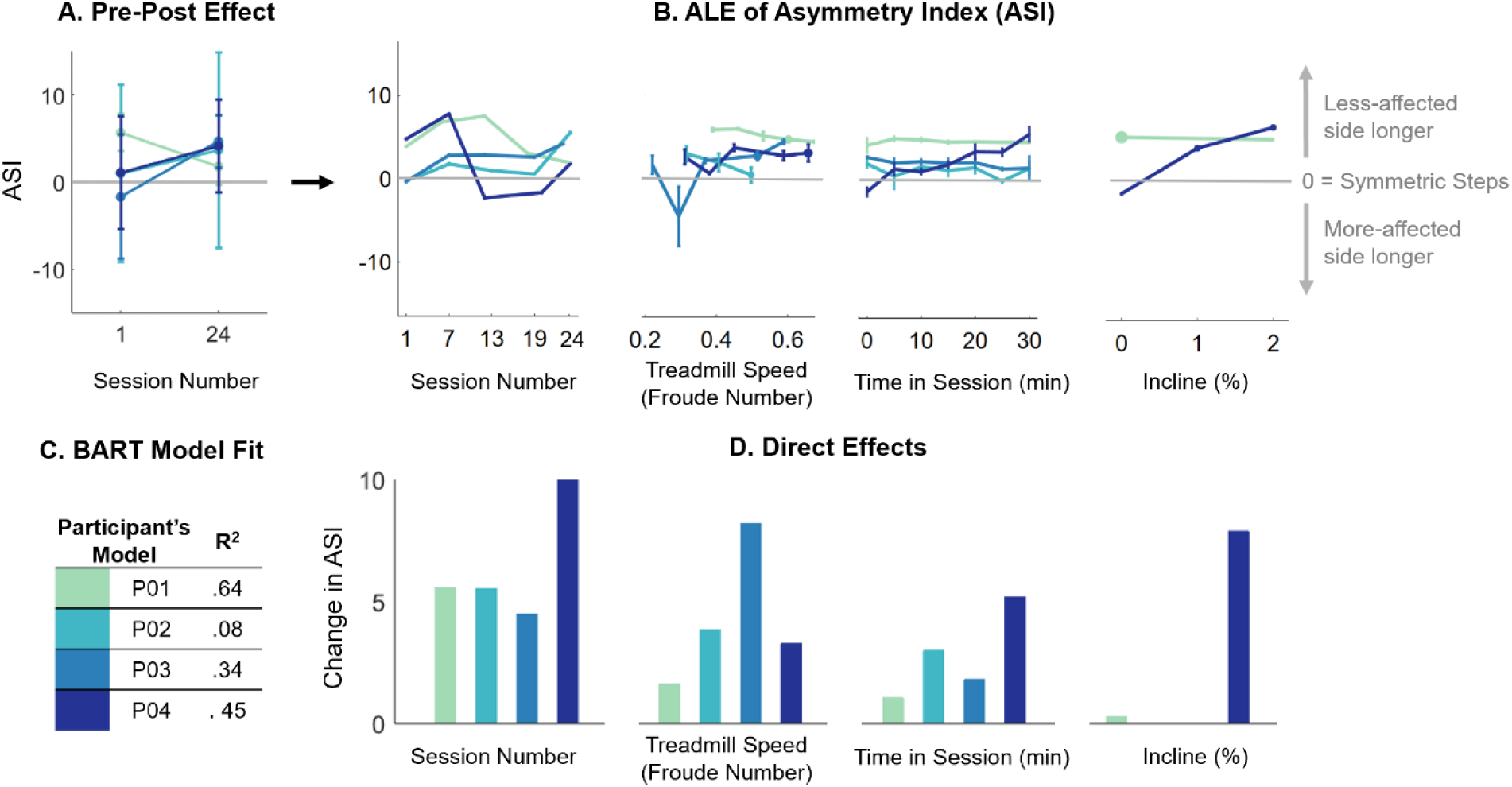
A) Pre-post effect of SBLTT on step length Asymmetry Index (ASI). B) BART results quantify direct effects of SBLTT on ASI. ALE plots for each input variable show the effect of that variable on ASI including session number, time within session, treadmill speed (Froude number), and treadmill incline. The size of the data point on each ALE plot depicts the relative number of data points in each bin. C) BART model fit (R^2^) for each participant. D) Direct effects of each input variable on the response variable, ASI, calculated from the change in the ALE plots in B).

### Impact of SBLTT on Step Width

Minimal changes were also observed in step width after SBLTT. The pre-post effects were highly variable between participants with an average increase in step width of 2.2 ± 23% (Figure 4A). BART models had poor to moderate fit for explaining step width variability (0.06 < R^2^ < 0.63; Figure 4C). SBLTT session number had a direct effect of increasing step width for two participants (P02: 11%; P03: 29%) and decreasing step width for two participants (P01: 56%; P04: 30%; Figure 4B and D). The direct effects from BART indicated minimal effects of treadmill speed, treadmill incline, or time in session on step width (direct effect < 0.03 m/m).

**Figure 4.**
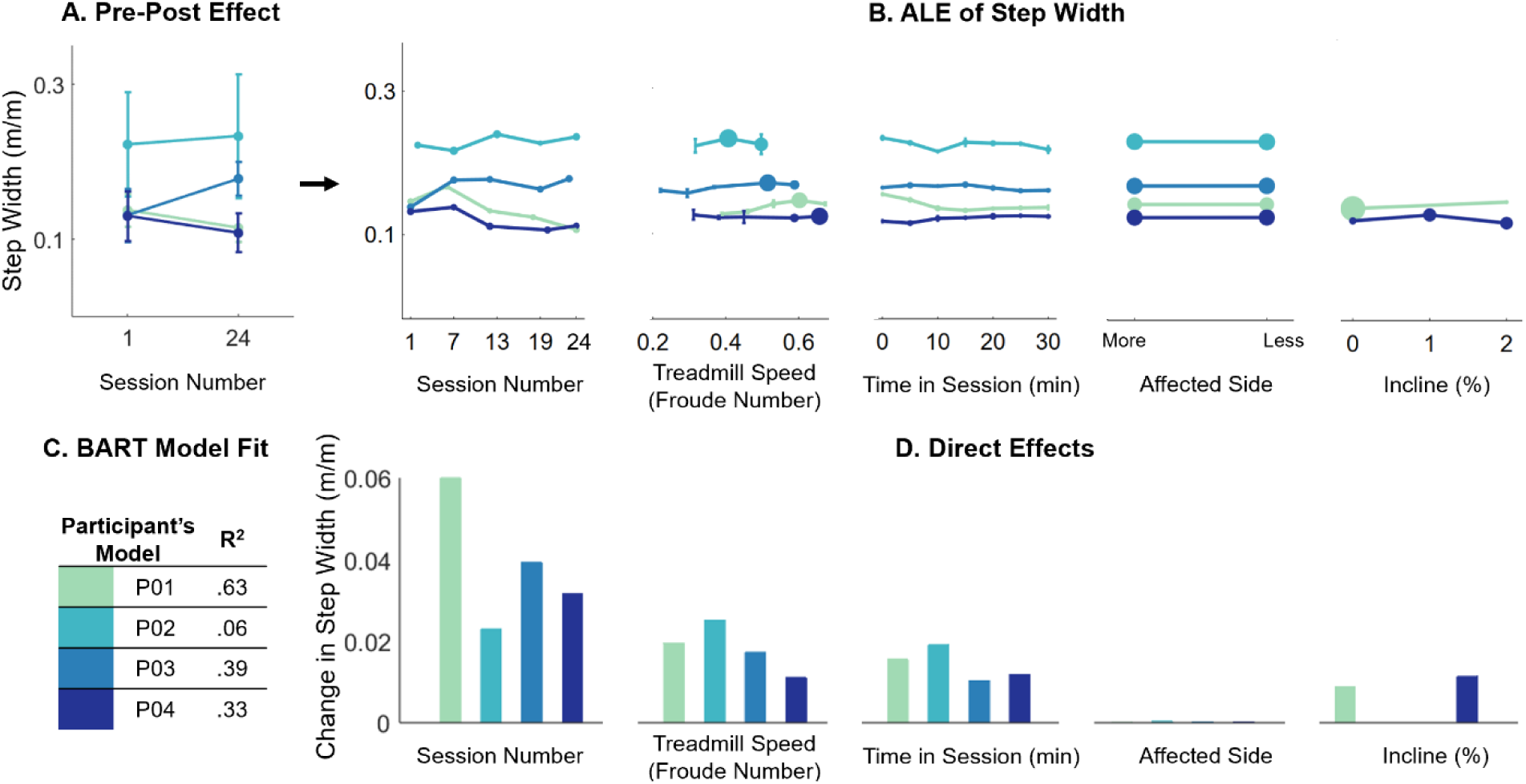
A) Pre-post effect of SBLTT on step width when the more affected side is ahead. B) BART results quantify direct effects of SBLTT on step width. ALE plots for each input variable show the effect of that variable on step width including session number, treadmill speed (Froude number), time within session, side, and treadmill incline. The size of the data point on each ALE plot depicts the relative number of data points in each bin. C) BART model fit (R^2^) for each participant. D) Direct effects of each input variable on the response variable, step width, calculated from the change in the ALE plots in B).

### In Silico: BART Accuracy

Evaluation of 150 virtual participants demonstrated that BART could accurately identify specified changes in step length across sessions. The specified and predicted step length progression with SBLTT were correlated across virtual participants (R^2^ = 0.73; Figure 5A). Greater step-to-step variability reduced BART model fit (R^2^ = 0.70, Figure 5B). However, the magnitude of step-to-step variability in a virtual participant’s data had no relationship with BART prediction accuracy (R^2^ = 0.01, Figure 5C). When step length progression was set to plateau, BART models were also able to identify specified plateau points (R^2^ = 0.87; Figure 5D).

**Figure 5.**
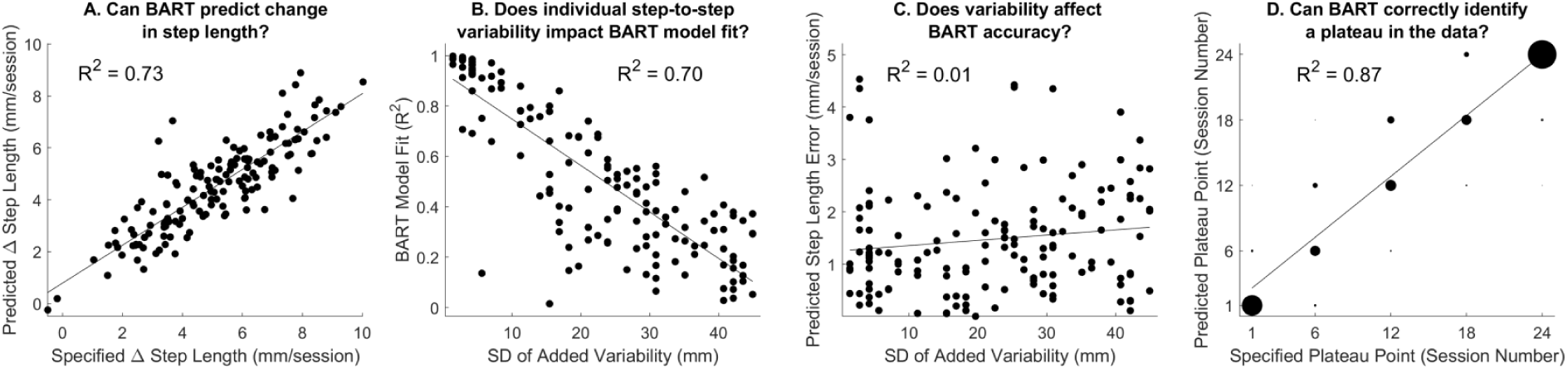
Results from *in silico* methods: A) Specified and predicted change in step length between sessions (mm/session) for virtual participants were highly correlated, R^2^ = 0.73. B) Increasing step-to-step variability of virtual participants decreased the variance explained by BART models, R^2^ = 0.70. C) Increasing step-to-step variability did not affect the accuracy of BART, R^2^ = 0.01. D) When change in step length was halted after a specified session – defined as the plateau point – the BART models of virtual participants accurately identified the plateau point, R^2^ = 0.87. Given the overlap in points for identifying plateaus, the size of each data point corresponds to the number of virtual participants in that bin.

## Discussion

We demonstrated that 24 sessions of SBLTT had a pre-post effect of increasing step length in four children with CP. Additionally, after controlling for other factors, SBLTT had a direct effect on increasing step lengths in three participants. In contrast, there were minimal pre-post or direct effects of SBLTT on step length asymmetry and step width. We further demonstrated that BART can be used to accurately quantify the direct effects of gait training using *in silico* datasets that mimicked the step-to-step variability and training progression expected across sessions.

Children with CP often exhibit reduced walking speed and step length compared to typically developing (TD) peers [23–25]. Reduced step lengths require a higher cadence to achieve a given walking speed, which increases metabolic cost of walking and negatively impacts mobility [26]. Altered speed modulation strategies in CP may reflect changes in neuromuscular control, musculoskeletal physiology, spasticity, contracture, or skeletal alignment [27–30]. We demonstrated that SBLTT had a direct effect on increasing step length after controlling for covariates of treadmill speed, incline, side of the body, and time in session. All participants, except for P02’s direct effect of SBLTT on step length, surpassed the minimum clinically important difference (MCID) for a large effect of SBLTT on increased step length from both the pre-post and direct effects of SBLTT (6.7 % and 6.3 % for GMFCS I and II, respectively) [31]. However, it remains unclear by what mechanisms SBLTT improved step length, i.e. changes in motor control, spasticity, or dynamic muscle lengthening.

Children with CP often exhibit increased step length asymmetry and width compared to their TD peers, indicators of reduced walking stability [23,24,32]. SBLTT had minimal effects on step length asymmetry and step width. During all SBLTT sessions, children were encouraged to use the handrails for safety, which likely influenced these spatiotemporal outcomes as the handrail offered an additional point of contact for stability [33]. While we aimed to keep handrail usage consistent across sessions, we did not record whether usage differed within or between participants.

The BART models provided excellent fit for step length among the older participants but explained less of the variance in step length asymmetry and step width. This suggests that there may be additional variables needed in our causal model for asymmetry and step width to capture the factors influencing these outcomes during SBLTT such as handrail usage, perceived fatigue, audiovisual input (*e.g.,* verbal cues), or trunk movement [34,35]. For the youngest participant (P02), all three models had poor fit (R^2^ < 0.21). Our *in silico* analysis demonstrated this is likely driven by the highly variable gait pattern of this participant, which is characteristic of this age range due to developing motor and postural control [36,37]. However, the *in silico* results also indicated that increased variability did not negatively affect the accuracy of the ALE plots (Figure 5C). While the BART models could explain variation in step length for the older participants with more mature walking, other factors may need to be included in the model for younger children, such as neuromuscular activity or engagement.

Causal modeling and machine learning, especially methods that are nonlinear and interpretable like BART, are useful tools to parse complex, individualized rehabilitation responses. One limitation is that these methods do require a large amount of data – in this case, many steps monitored during treadmill training. Applying these methods to other environments where there may be limitations to acquiring large datasets may require the use of emergent techniques in wearable sensing. With only four participants, the generalizability of these results to other children with CP is limited. However, they provide a framework for a more detailed evaluation of individual intervention progression for highly heterogeneous populations like children with CP. Further, we only recorded 5 of 24 sessions to track progression, but quantifying spatiotemporal outcomes at each session may give further insight into treatment progression. Despite these limitations, this work shows the potential efficacy of using BART for quantifying individual responses to interventions, especially for tracking training-specific parameters, like session length or treadmill speed and incline, that can be adjusted to personalize rehabilitation. There may also be additional clinical information not included here to consider in the causal model to capture factors important for SBLTT responses, such as changes in spasticity, range of motion, or motor control between sessions. Future research to integrate causal models of altered neuromuscular physiology with models of rehabilitation protocols represents an exciting area to further understand individual differences in intervention responses.

Using BART, we quantified step-by-step personalized response to SBLTT and demonstrated that SBLTT had a direct effect on increasing step length in three children with CP, even after controlling for increases in walking speed and other training parameters. These methods can be expanded to evaluate other gait training programs and other populations, such as stroke or Parkinson’s disease. Further, using wearable sensors or video-based techniques could offer greater insights on step-by-step changes in gait during rehabilitation [38–40]. Pairing these rich data sources with causal modeling and machine learning paradigms will help to personalize and optimize rehabilitation and enhance mobility for people with disabilities.

## Data Availability

All data produced in the present work are contained in the manuscript.

## Acknowledgements

The researchers would like to thank the families and research participants for their time and energy dedicated to this research and Dr. Cristine Agresta for use of her pressure instrumented treadmill. This work was supported by Seattle Children’s Hospital CP Research Pilot Study Fund 2020 Award, UW Rehabilitation Medicine Walter C, and Anita C. Stolov 2021 Research Fund, and NSF Graduate Research Fellowship Program Award [DGE-1762114].

